# Exposure to public housing reverses the association between neighborhood disadvantage and eosinophilic inflammation in patients with sickle cell disease

**DOI:** 10.1101/2025.01.27.25321223

**Authors:** Sarah McCuskee, Yueh-Hsiu Mathilda Chiu, Mariel McCann, Rosalind J Wright, Jeffrey A Glassberg

## Abstract

Sickle cell disease (SCD) is a single-gene illness characterized by chronic inflammation, decreased quality of life, and early mortality; however, outcomes are highly variable between individuals, suggesting a substantial role of social and environmental factors in disease outcomes. Data suggest that individuals living with SCD have greater eosinophil counts and activation, and higher prevalence of asthma and wheezing than those without SCD, suggesting a role for eosinophilic inflammation in SCD. In other diseases, eosinophilic inflammation has been linked to social and environmental factors, particularly in minoritized populations. To date, however, few human studies have explored the pathophysiology of social and environmental exposures in SCD. This study tested whether eosinophilic inflammation was related to location-based measures of social disadvantage or public housing in 79 individuals with SCD, without diagnoses of asthma, who were prospectively followed over one year. Home addresses were geocoded and matched to principal-components derived, validated measures of local neighborhood social disadvantage and to locations of public housing facilities. Serum peripheral eosinophils, IL-13 and IL-5 were measured every 8 weeks. In fully-adjusted models, statistically significant, linear relationships were observed between the degree of social disadvantage and level of eosinophilic inflammation; however, the direction of that relationship was opposite for patients who live in public housing and those who do not. For those living in public housing, greater social disadvantage was associated with increased eosinophilic inflammation. For those in private housing, greater social disadvantage was associated with progressively less eosinophilic inflammation. These strong, and somewhat unexpected, relationships demonstrate that subtle differences in social exposures and home environment have differential effects on inflammatory profiles which may have larger implications for disease and health, especially in chronic diseases such as SCD. To understand the mechanisms of these effects may require highly granular studies that catalog the many factors underlying the social environment and inflammation.

## 1. Introduction

Social drivers related to where individuals live and the resources they can access are known to impact health; however, postulated mechanisms have mostly referenced behavioral mediators (for either clinicians or patients), which then influence health through established pathways such as smoking, diet, attendance at appointments, or adherence to medications. Behavioral interventions are thus commonly prioritized for highly exposed or under-resourced individuals and communities.^1^ However, an alternate hypothesis is that social drivers directly cause biochemical changes, acting through changes in cytokine and chemokine signaling which directly affect health outcomes (and possibly behaviors).^2^ In this paradigm, effective interventions might focus on early identification and mitigation of structural social and environmental stressors to prevent morbidity and mortality, instead of behavioral adaptation to structural stressors and treatment of illness once it develops.

Many patients with sickle cell disease (SCD) are members of under-resourced communities that are often involuntarily exposed to elevated levels of environmental stressors. SCD is a genetic disparity illness caused by single-gene mutations in the beta-globin gene, which causes conformational changes in hemoglobin affecting polymerization and causing red blood cells to “sickle” when under stresses including deoxygenation or dehydration.^3^ Approximately 90% of individuals living with SCD in the United States identify as Black or African American.^4^ However, despite its single-gene etiology, outcomes are highly variable between individuals, suggesting that the role of social and environmental drivers in disease outcomes is substantial.^5^ The role of inflammation in pathophysiology is critical; however, scientific understanding of the multiple inflammatory processes ongoing in SCD is relatively nascent.

In particular, eosinophils, shown to exist in greater numbers and more activated states in people living with SCD than in controls^6^, are thought to drive the production of reactive oxygen species^7^, and decrease in number and adhesive properties with hydroxyurea therapy^6^ (hydroxyurea decreases mortality in patients with SCD^8,9^), indicating importance in pathophysiology. This eosinophilic inflammation may also contribute to the high prevalence of asthma (17-48%) in SCD^10^. Having both asthma and SCD predicts poor outcomes^11^; even individuals living with SCD without asthma who have pulmonary inflammation indicated by wheeze or cough have increased complications including hospitalization and acute chest syndrome.^12^ However, mechanisms driving eosinophilic inflammation in SCD are very poorly understood.

In this study, we tested the hypothesis that exposure to hyperlocal neighborhood disadvantage impacts eosinophilic inflammatory signaling and clinical outcomes in patients with SCD, and investigated exposure to public housing as an effect modifier of this relationship.

## 2. Materials and Methods

### 2.1. Study design

This is a prospective longitudinal observational study using data from the IMPROVE2 randomized clinical trial, which included patients with sickle cell disease (HbSS or HbS β-thal 0) who were on maximal medical therapy (hydroxyurea or ineligible for hydroxyurea) and who had episodic coughing or wheezing. Individuals with diagnoses of asthma or who screened positive for potentially undiagnosed asthma were excluded using a standardized algorithm^13,14^. Patients were assigned to either placebo or treatment with inhaled mometasone (220mcg daily, weeks 0-48) and followed every eight weeks over 1 year with serial laboratory and cytokine measurements. Full methodology for the trial is available separately (NCT03758950). Briefly, 80 participants living with SCD in New York City and the surrounding communities were enrolled from December 2018 to May 2022 and 79 had at least one blood draw for outcomes as below, making them eligible for inclusion in this study; in total, 272 participant-timepoints were studied. The study was approved by the Institutional Review Board.

### 2.2. Outcome data

Complete blood count and differential were assessed. Serum for eosinophil-related inflammatory markers (IL-5, IL-13) was collected in Vacutainer serum separator tubes (BD, Franklin Lakes, NJ, USA), and stored on ice for less than 4 hours before centrifugation, then separated and stored at -80°C until analysis. Cytokines were measured using multiplexed real-time quantitative PCR (O-Link Inflammation proteomics platform, O-link, Waltham, MA, USA), with all samples run in the same batch and randomized within plates, and data were intensity-normalized prior to analysis, producing normalized protein expression (NPX) values on the log2 scale. Samples were examined for outliers using principal components analysis in the R package OlinkAnalyze and one outlier was removed.

### 2.3. Exposure data

We geocoded participants’ home addresses and matched these to a census tract-level neighborhood disadvantage z-score. Specifically, a measure of neighborhood disadvantage was derived by linking enrollment addresses with aggregated data (census tract) from the 2015 US Census indexed as an average z-score for percentages of residents living below poverty, the unemployed, non-US citizens, non-white, less than college degree, and receiving public assistance in the neighborhood using previously accepted methods derived by Sampson et al^15,16^. Z-scores are interpreted in a relative sense; neighborhoods with the most positive z-scores are most disadvantaged and those at the most negative end are least disadvantaged. Census tracts are the smallest administrative subdivision provided by the United States Census and typically include approximately 4000 individuals. No participant changed home address during the study period. Geospatial processing was completed in ArcGIS Pro using the ArcGIS World Geocoding Service.

### 2.4. Covariates

Participants were screened with questionnaires by research staff at enrollment to determine gender, medication status, baseline symptom burden, and other clinical history.

We created an indicator variable for public housing exposure based on the New York City Housing Authority public housing developments database, and defined that a participant living at home addresses within one city block (80 meters) of a public housing as ‘exposed’ to public housing.^17^ Other covariates thought to represent confounders based on a Directed Acyclic Graph were included^18^, namely: age, gender, and use of hydroxyurea, which affects leukocyte differentiation. Because we used data from a clinical trial, we also adjusted for assignment to inhaled mometasone in the trial.

### 2.5. Statistical analysis

The descriptive statistics of the outcomes (IL-13, IL-5, and relative/absolute eosinophils), social disadvantage z-score, and covariates were summarized by public housing exposure status. To examine the association between neighborhood disadvantage and the outcomes, mixed effects linear regressions by maximum likelihood with a random intercept per patient to allow for longitudinal repeated measures were conducted using the *nlme* package in R^19^. To eliminate omitted variable error in modeling of the random intercepts resulting from the correlation between the grouping variable (participant) and predictors (residence-related exposures per participant), we applied Bafumi and Gelman’s method, including the mean participant-level predictors in the model.^20^ Exposure to public housing was investigated as an effect modifier of this relationship, using both stratified analyses and interaction models. Model diagnostics were performed, and sensitivity to distribution was assessed using Bayesian multilevel robust linear regression with Gaussian and Student’s distributions in R package *brms*^21^. All statistical analyses were conducted in R 4.1.1.

## 3. Results and Discussion

Descriptive statistics are presented in Table 1. Covariates were broadly similar between individuals exposed to public housing or not exposed, including age, gender, hydroxyurea use, and exposure to inhaled mometasone. Predictors and outcomes were not similar between groups.

**Table 1:** Descriptive statistics for patient cohort, stratified by residence in public housing.

|  | Public Housing Residence |  |  |
| --- | --- | --- | --- |
|  | No<br>(N=59) | Yes<br>(N=20) | Overall<br>(N=79) |
| Neighborhood Disadvantage (z-score) |  |  |  |
| Mean (SD) | 0.555 (0.647) | 0.795 (0.662) | 0.623 (0.655) |
| Missing | 11 (18.6%) | 1 (5.0%) | 12 (15.2%) |
| Age |  |  |  |
| Median [IQR] | 31.0 [24, 40.5] | 30.5 [28, 35] | 31.0 [25.8, 39] |
| Gender |  |  |  |
| Male | 26 (44.1%) | 10 (50.0%) | 36 (45.6%) |
| Female | 33 (55.9%) | 10 (50.0%) | 43 (54.4%) |
| Hydroxyurea use |  |  |  |
| No | 15 (25.4%) | 5 (25.0%) | 20 (25.3%) |
| Yes | 43 (72.9%) | 15 (75.0%) | 58 (73.4%) |
| Missing | 1 (1.7%) | 0 (0%) | 1 (1.3%) |
| Treatment group |  |  |  |
| Control | 27 (45.8%) | 10 (50.0%) | 37 (46.8%) |
| Mometasone | 27 (45.8%) | 8 (40.0%) | 35 (44.3%) |
| Missing | 5 (8.5%) | 2 (10.0%) | 7 (8.9%) |
| IL-13 |  |  |  |
| Mean (SD) | -0.987 (0.595) | -0.897 (0.555) | -0.963 (0.585) |
| IL-5 |  |  |  |
| Mean (SD) | -1.15 (0.660) | -0.524 (1.89) | -0.985 (1.16) |
| Relative eosinophils (% leukocytes) |  |  |  |
| Mean (SD) | 3.03 (4.65) | 2.18 (2.06) | 2.80 (4.14) |

After adjustment for gender, age, exposure to inhaled mometasone, and current hydroxyurea use, there was a significant, inverse relationship between the census tract level of neighborhood disadvantage and eosinophilic inflammation (IL-13 level and eosinophil percent) in this population of individuals living with SCD. IL-13, an inflammatory mediator linked to allergic inflammation, declined with higher neighborhood disadvantage (b = -0.38, 95% confidence interval -0.64 – -0.11). Relative eosinophilia (as proportion of total leukocytes) in peripheral blood was also declined in participants living in more disadvantaged neighborhood (b = -1.14, 95%CI -2.14 – - 0.14). However, the relationship between neighborhood disadvantage and relative eosinophilia was modified by exposure to public housing. In individuals exposed to public housing, elevated neighborhood disadvantage z-score was associated with increased levels of eosinophilia, as demonstrated in Figure 1 and Table 2 (*p* for interaction = 0.02). These models had appropriately distributed residuals with acceptable homoscedasticity; Bayesian multilevel robust linear regression using Student’s distribution and noninformative priors produced similar results for all models.

**Figure 1.**
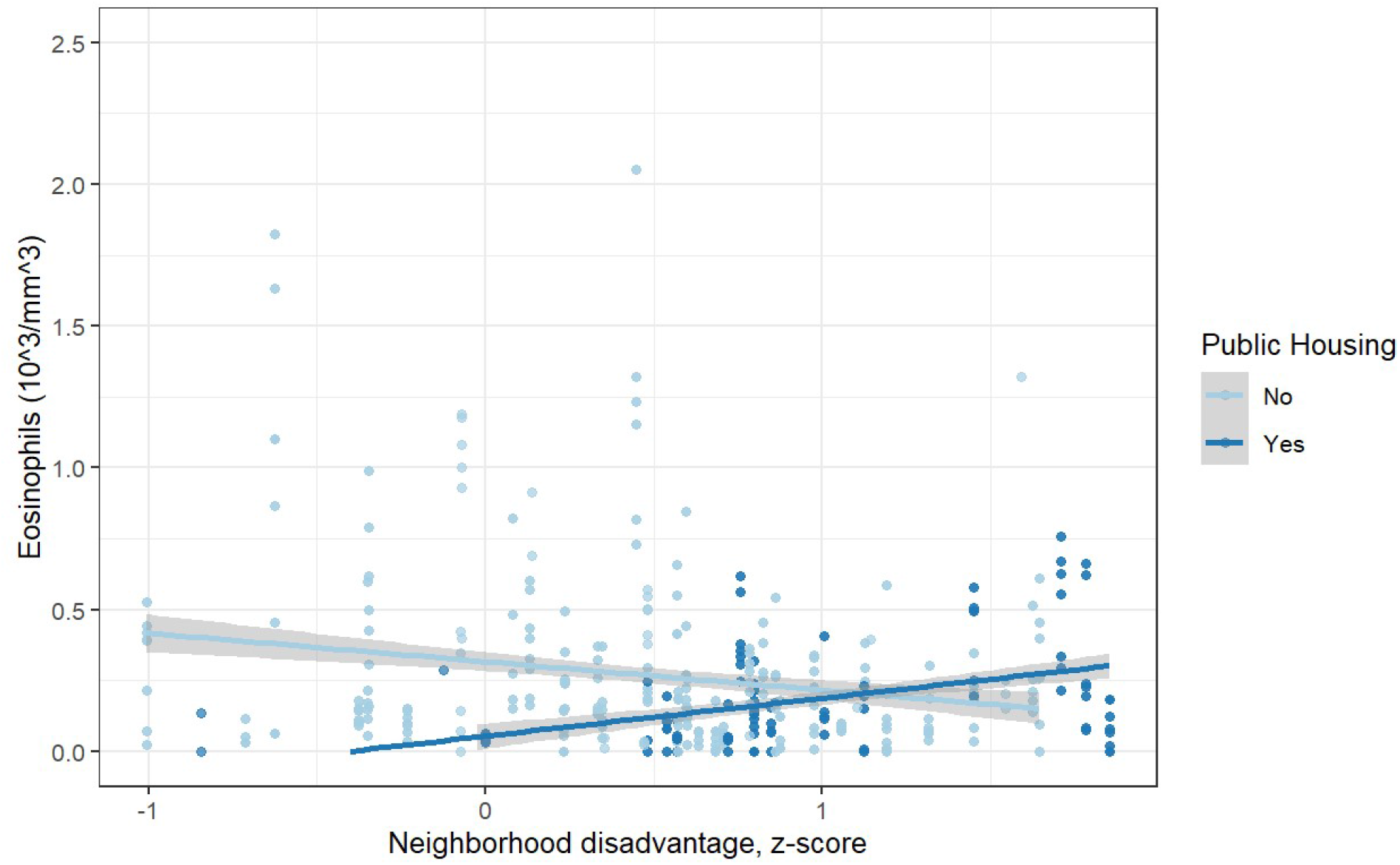
Peripheral eosinophilia and increasing neighborhood disadvantage, stratified by residence in public housing. Shading indicates 95%CI.

**Table 2.** Longitudinal mixed effects linear models with random intercept by patient, adjusting for sex, age, trial treatment assignment to inhaled mometasone, and current hydroxyurea use.

| *Predictors* | IL-13 |  |  | IL-5 |  |  | Eosinophils |  |  |
| --- | --- | --- | --- | --- | --- | --- | --- | --- | --- |
|  | *Estimate* | *CI* | *p* | *Estimate* | *CI* | *p* | *Estimate* | *CI* | *p* |
| Neighborhood Disadvantage (z-score) | -0.30 | -0.53 – -0.07 | **0.012** | -0.14 | -0.67 – 0.39 | 0.610 | -1.14 | -2.14 – -0.14 | **0.026** |
| Public Housing Exposure | 0.27 | -0.03 – 0.58 | 0.080 | 0.75 | 0.04 – 1.46 | **0.040** | -2.26 | -4.20 – -0.32 | **0.024** |
| Participant age at enrollment | 0.00 | -0.01 – 0.02 | 0.752 | 0.01 | -0.02 – 0.05 | 0.470 | -0.00 | -0.06 – 0.05 | 0.927 |
| Gender: Female | -0.07 | -0.36 – 0.23 | 0.660 | 0.27 | -0.42 – 0.95 | 0.439 | -0.39 | -1.52 – 0.73 | 0.487 |
| Mometasone exposure | 0.07 | -0.22 – 0.36 | 0.621 | 0.36 | -0.31 – 1.03 | 0.283 | -0.03 | -1.16 – 1.10 | 0.960 |
| Hydroxyurea use | 0.12 | -0.21 – 0.45 | 0.471 | 0.18 | -0.59 – 0.94 | 0.643 | -0.45 | -1.70 – 0.80 | 0.477 |
| (Public Housing) * (Neighborhood Disadvantage z-score) interaction term |  |  |  |  |  |  | 2.27 | 0.35 – 4.18 | **0.021** |
| **Random Effects** |  |  |  |  |  |  |  |  |  |
| $\sigma^2$ | 0.11 | | | 0.15 | | | 3.52 | | |
| $T_{00}$ | 0.24<sub>ptid</sub> | | | 1.40<sub>ptid</sub> | | | 2.98<sub>ptid</sub> | | |
| ICC | 0.69 |  |  | 0.90 |  |  | 0.46 |  |  |
| Patients | 59<sub>ptid</sub> |  |  | 59<sub>ptid</sub> |  |  | 59<sub>ptid</sub> |  |  |
| Patient-visits | 272 |  |  | 272 |  |  | 268 |  |  |
| Conditional R<sup>2</sup> | 0.725 |  |  | 0.912 |  |  | 0.516 |  |  |

Epidemiologic studies have demonstrated that allergic asthma is disproportionately prevalent in Black/African American and Latinx/Hispanic-identifying communities, and that exposure to poor housing quality, in particular exposure to public housing with cockroaches, rats, and water leaks, is a risk factor for asthma.^1,6^ However, existing literature has not fully elucidated the biochemical changes or causal mechanisms implicated in these ecological associations. In addition, eosinophilic inflammation in individuals without asthma is relatively poorly studied, and this study is the first to explore mechanistic drivers of eosinophilic inflammation in non-asthmatic patients with SCD. Augmentations in eosinophil responsiveness in SCD have been previously suggested in small *in vitro* studies^22^. However, the biology of allergic and eosinophilic inflammation in SCD is very poorly understood. Drivers of allergic inflammation such as disadvantage and housing type have not previously been explored in this population.

The finding that IL-5 is positively associated with exposure to public housing (with a similar statistical trend for IL-13), while IL-13 is negatively associated with neighborhood disadvantage, suggests that eosinophil chemotaxis (of which IL-5 is an important peripheral mediator) may be altered based on social and environmental stressors. Eosinophils express IL-13 in lung tissue when activated, but this is independent of peripheral circulating eosinophilia or IL-13 in mouse models^23^; thus the opposing effects of IL-5 vs. IL-13 and peripheral eosinophilia in our population may reflect peripheral vs. lung-specific inflammatory responses. Further research should explore tissue-specific or sputum inflammatory responses related to environmental triggers.

The negative effect of public housing on eosinophilia persisted when examining the outcome of absolute eosinophil count (b = -0.26, 95% CI -0.51 – -0.01), but neighborhood disadvantage was not significantly associated with absolute eosinophilia. We modeled relative eosinophilia as an outcome in patients with SCD because many factors affect the overall leukocyte count including medications and exposure to infectious diseases; differential distribution of these unmeasured covariates may account for the differences between models predicting relative eosinophilia and absolute eosinophilia. However, the importance of exposure to public housing persists across both models.

This study’s main limitation was inability to ascertain social disadvantage z-scores for patients residing outside of New York state, leading to some missing data (15%). Covariates except for trial treatment assignment were similar in participants with or without social disadvantage z-scores; more participants with missing data for neighborhood disadvantage were assigned to inhaled steroids. Due to differences by outcome in trial treatment assignment, these values were not imputed. This suggests that future work with a larger sample size should include more detailed ascertainment of social disadvantage and explore the generalizability of these results outside of NY state, although given the differing results, exposures may overall differ substantially in this subpopulation. However, to our knowledge this is the first mechanistic exploration of the impact of social disadvantage and housing environment in patients with SCD. The study’s strengths include its longitudinal outcomes and its inclusion of symptomatic patients (with cough or wheeze), as well as its rigorous exclusion of individuals with asthma, which allowed mechanistic exploration of allergic inflammatory pathways in SCD itself.

## 4. Implications

Studying the impact of social and environmental factors within a majority Black/African American-identifying population avoids the use of “race” as an explanatory factor for health disparities. This allows identification of the true and actionable social and environmental drivers, and their mechanisms, within populations that are racially/ethnically minoritized in the United States. In this study, we uncover associations between eosinophilic inflammation and chemotaxis and neighborhood disadvantage in patients with SCD, which we hypothesize may reflect differential chemotaxis of eosinophilic inflammatory cells. The reversal of this relationship in individuals exposed to public housing may signify important mechanistic effects of allergic inflammation based on housing characteristics or stress exposures, and should be further investigated to begin to mitigate the structural drivers of eosinophilic inflammation.

## Data Availability

All data produced in the present study are available upon reasonable request to the authors, except as limited by applicable privacy and institutional policies.

